# Molecular malaria surveillance using a novel protocol for extraction and analysis of nucleic acids retained on used rapid diagnostic tests

**DOI:** 10.1101/2020.02.17.20023960

**Authors:** Etienne A. Guirou, Tobias Schindler, Salome Hosch, Olivier Tresor Donfack, Charlene Aya Yoboue, Silvan Krähenbühl, Anna Deal, Glenda Cosi, Linda Gondwe, Grace Mwangoka, Heavenlight Masuki, Nahya Salim, Maxmillian Mpina, Jongo Said, Salim Abdulla, Stephen L. Hoffman, Bonifacio Manguire Nlavo, Carl Maas, Carlos Cortes Falla, Wonder P. Phiri, Guillermo A. Garcia, Marcel Tanner, Claudia Daubenberger

**Affiliations:** Department of Medical Parasitology and Infection Biology, Swiss Tropical and Public Health Institute, Basel, Switzerland; University of Basel, Basel, Switzerland; Medical Care Development International, Malabo, Equatorial Guinea; Ifakara Health Institute, Bagamoyo Branch, United Republic of Tanzania; Department of Paediatrics and Child Health, Muhimbili University of Health and Allied Sciences, Dar Es Salaam, Tanzania; Sanaria Inc., Rockville, Maryland, USA; Marathon EG Production Ltd, Malabo, Equatorial Guinea

**Keywords:** Nucleic Acid Extraction, Molecular Malaria Surveillance, Rapid Diagnostic Test (RDT), Reverse Transcription Quantitative Polymerase Chain Reaction (RT-qPCR), Artemisinin resistance, kelch 13

## Abstract

The use of malaria rapid diagnostic tests (RDTs) as a source for nucleic acids that can be analyzed via nucleic acid amplification techniques has several advantages, including minimal amounts of blood, sample collection, simplified storage and shipping conditions at room temperature. We have systematically developed and extensively evaluated a procedure to extract total nucleic acids from used malaria RDTs. The co-extraction of DNA and RNA molecules from small volumes of dried blood retained on the RDTs allows detection and quantification of *P. falciparum* parasites from asymptomatic patients with parasite densities as low as 1 Pf/µL blood using reverse transcription quantitative PCR. Based on the extraction protocol we have developed the ENAR (Extraction of Nucleic Acids from RDTs) approach; a complete workflow for large-scale molecular malaria surveillance. Using RDTs collected during a malaria indicator survey we demonstrated that ENAR provides a powerful tool to analyze nucleic acids from thousands of RDTs in a standardized and high-throughput manner. We found several, known and new, non-synonymous single nucleotide polymorphisms in the propeller region of the kelch 13 gene among isolates circulating on Bioko Island, Equatorial Guinea.

## Introduction

Malaria remains a global public health issue with an estimated 228 million cases resulting in an estimated 405,000 deaths in 2018^1^. *P. falciparum* (*Pf*) is the most pathogenic malaria species accounting for the vast majority of malaria cases and deaths. Malaria surveillance, the continuous and systematic collection, analysis and interpretation of epidemiological data, is the core monitoring and evaluation tool for malaria control programs, and provides the framework for effective allocation of resources^2^. A critical surveillance measure, which closely reflects malaria transmission intensity, is the parasite rate; the proportion of the population found to carry parasites in their peripheral blood^3,4^. Malaria rapid diagnostic tests (RDTs) are the most widely used technique to measure parasite rates in endemic countries. In sub-Saharan Africa, RDTs have almost completely replaced light microscopy for malaria diagnosis, with an estimated 75% of malaria tests conducted using RDTs in 2017^1^. RDTs are relatively low cost, provide fast result turnaround time, are widely available and easy to use. However, there are also disadvantages including low sensitivity, resulting in poor performance among asymptomatic individuals^5^ and the widespread emergence of *pfhrp2* deletions in certain regions^6^ whereby RDTs fail to detect malaria infection.

Nucleic amplification techniques (NATs), such as polymerase chain reaction (PCR), not only show higher sensitivities than RDTs^5,7^ but also allow further characterization of *Pf* isolates using molecular markers. Surveillance of drug-resistant *Pf* strains, based on analysis of resistance-associated molecular markers, is a widely used and valuable epidemiological tool^8^. In sub-Saharan Africa, malaria treatment relies heavily on artemisinin-based combination therapy (ACT). The implementation of surveillance programs for early detection of emerging artemisinin-resistant *Pf* strains will be the key to prevent the spread across the continent^9^. Artemisinin-resistant *Pf* strains were first reported in Cambodia^10,11^ and remain a public health concern in South East Asia but have not yet been found to be widespread in Africa, South America or Oceania^12^. Non-synonymous mutations in the propeller region of the *Pf* kelch 13 gene (pfk13) were discovered as molecular markers for artemisinin resistance^13^.

Residual blood from RDTs are an ideal source for nucleic acids (NAs) to be used for NAT-based resistance markers screening and present several advantages, including simplicity and cost-effectiveness of sample collection, as well as simplified storage and shipping conditions at room temperature (RT). Over the past decade, several reports have been published describing the use of DNA extracted from used RDTs for molecular analysis of malaria parasites (studies summarized in Supplementary Table S1)^14–24^. However, most studies that tried to address the question of using RDTs as source of DNA were conducted with small sample sizes and focused on demonstrating the feasibility of extracting DNA rather than fitting this approach for molecular surveillance of malaria at larger scale. We identified three key areas that are critical to develop a surveillance tool based on molecular analysis of used RDTs: i) accessing a representative collection of RDTs and creating an effective selection and sorting strategies for RDTs of interest. ii) high-throughput extraction and analysis of NAs from RDTs with minimal hands-on time and focus on reproducibility and quality control throughout the entire extraction process. iii) increasing recovery of *Pf* NAs during the extraction process in order to include asymptomatic individuals with low parasite density infections. This manuscript outlines an overall strategy and the protocols for collecting, sorting and processing RDTs to extract the retained NA at large-scale in order to screen for single nucleotide polymorphisms (SNPs) in an artemisinin-resistance molecular marker in a dataset of thousands of healthy, malaria asymptomatic individuals. We systematically developed and extensively evaluated a procedure to extract NA from RDT. The “Extraction of Nucleic Acids from RDTs” (referred to as ENAR) approach is supported by custom-made software solutions that allow the analysis of thousands of RDTs in a standardized, reproducible and high-throughput manner.

We developed the ENAR approach in Tanzania and implemented the ENAR approach within Bioko Island Malaria Elimination Project’s (BIMEP) 2018 malaria indicator survey (MIS) conducted on Bioko Island, Equatorial Guinea. BIMEP is an island-wide intervention resulting in a substantial reduction in malaria, achieving a reduction in parasitemia of around 75% over the past 15 years^25^. Despite these achievements, malaria transmission remains stable on Bioko for an number of reasons, and recently a *Pf* isolate of African origin with artemisinin-resistance, including a novel non-synonymous mutation in pfk13, was identified in a 43-year-old man returning to China from Equatorial Guinea^26^. This reality underlies the importance of incorporating molecular techniques as monitoring and evaluation tools in malaria control programming.

## Results

### Blood stored on RDTs is a source of *Pf* DNA

First, we conducted a literature search of reports describing the use of NA extracted from RDTs as input templates for NAT-based detection of malaria parasites (Supplementary Table S1). A total of 11 studies were published between 2006 and 2019. All studies were limited to the extraction of DNA and used a variety of different extraction methods. Most extraction protocols were based either on the Chelex method (n=7) or silica column-based DNA extraction kits (n=6). One study extracted DNA from the entire RDT strip, all other studies used only predefined fragments of the RDT strip. These previous studies demonstrated that *Pf* DNA can be recovered from RDTs and amplified by NATs. Several studies genotyped drug resistance associated markers using sanger or next generation sequencing.

As the majority of these studies extracted DNA from RDTs of febrile clinical malaria cases, indicating high parasite densities, we first conducted a study to test feasibility of detecting Pf DNA from RDTs of asymptomatic individuals. We employed RDTs collected in a malaria survey conducted among asymptomatic children from three primary schools in the Mkuranga district of Coastal Tanzania. DNA was extracted from 190 RDTs and *Pf* DNA was recovered from 90.8% (59/65) of PfHRP2-positive RDTs, from 100% (5/5) of PfHRP2/pLDH-positive RDTs and from 11.7% (14/120) negative RDTs (Fig 1A).

**Figure 1.**
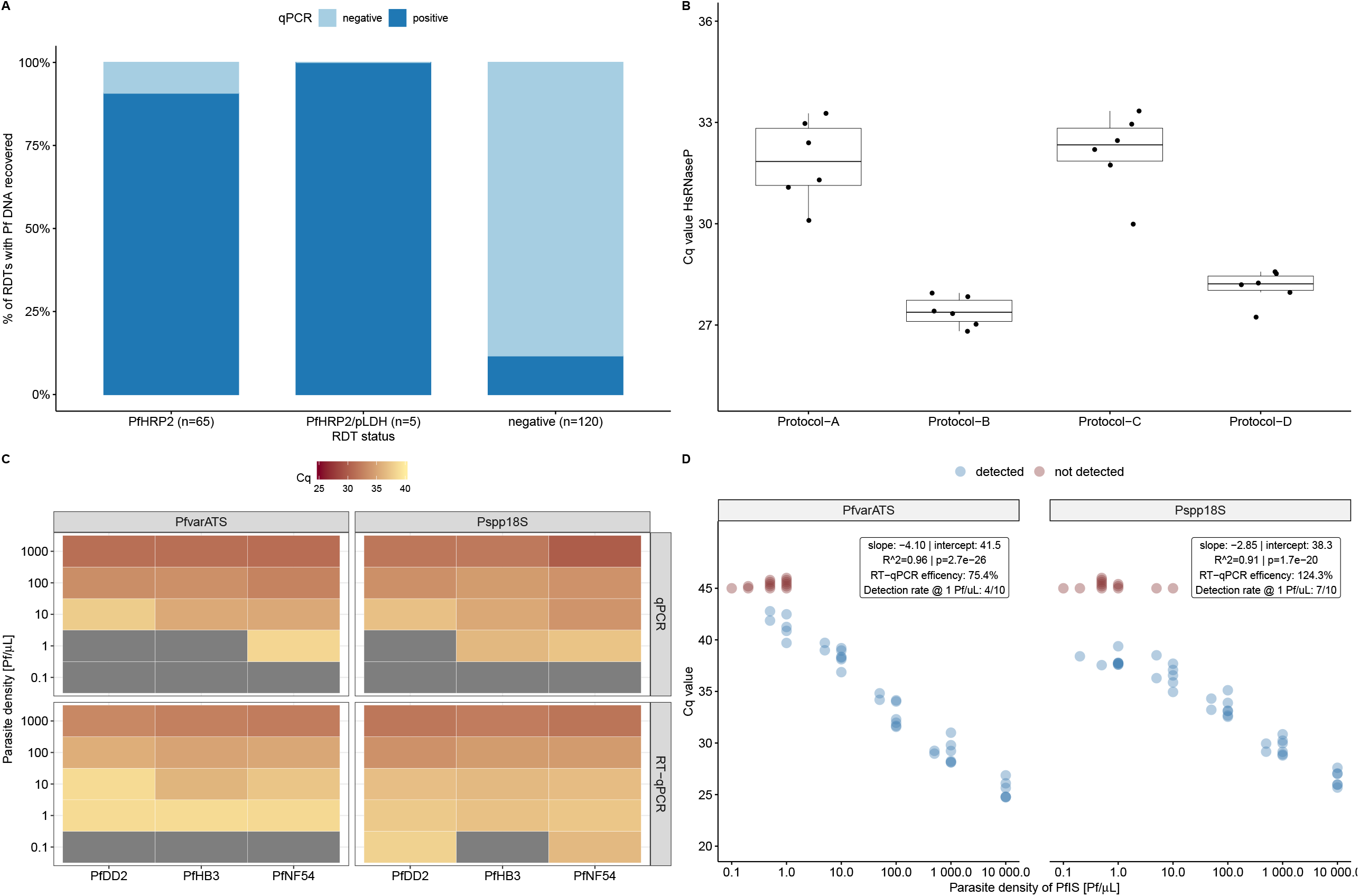
Extraction and detection of *Pf* NAs from used RDTs. A) Recovery rates of *Pf* DNA from RDTs collected in asymptomatic Tanzanian school children. B) Comparison of extraction performance of four protocols based on Cq values of the human *rnasep* gene. C) Association of parasite densities and Cq values of freshly prepared *Pf* strains (PfDD2, PfHB3 and PfNF54). Gray colour indicates failed detection. D) Correlation between parasite densities of serially diluted PfIS and Cq values for PlasQ targets. Red coloured dots represent samples where amplification failed.

Encouraged by the outcome of the school-based survey, we aimed to improve the extraction method from RDTs. As a proxy for the amount of extracted NAs, the Cq value of the human *rnasep* gene (HsRNaseP target), which is the internal control of the previously published PlasQ assay, was used to assess the overall performance of four different extraction procedures (Fig 1B). Side-by-side comparison of the four extraction procedures, named Protocol A through D, confirmed the superior performance of protocols B and D. Considering the costs and the fact that protocol D co-extracts RNA, we developed protocol D, which we renamed ENAR (Extraction of Nucleic Acids from RDTs). In order to identify the part of the RDT strip where most *Pf* NAs accumulate, we analyzed the sample pad (proximal part), the detection area (middle part), and the absorption pad (distal part) using ENAR. In RDTs probed with fresh blood, *Pf* NAs are found in all three parts, with more than 87% of the total extracted DNA conentrated in the middle part. RDTs spiked with frozen blood that is associated with red blood cell lysis resulted in an equal distribution of NA along the entire RDT strip (Supplementary Figure S1).

### Detection and quantification of *Pf* parasites based on ENAR protocol

We evaluated the ENAR protocol with cultured *Pf* strains including the strains PfDD2, PfHB3 and PfNF54 by preparing ten-fold serial dilutions in whole blood with parasite densities corresponding to 0.1 – 1000 Pf/µL. RDTs were spiked with 5 µL of diluted cultures, the NAs extracted by ENAR, and analyzed by qPCR and RT-qPCR (Fig 1C). Only the RT-qPCR assay resulted in detection of all three strains with the 1 Pf/µL parasite density. Furthermore, the Pspp18S-based RT-qPCR assay even detected two (PfDD2 and PfNF54) out of the three *Pf* strains at a concentration of 0.1 Pf/µL. This result demonstrates that the ENAR clearly co-extracts DNA and RNA. The *Pf* 18S ribosomal RNA, detected by the Pspp18S RT-qPCR assay, is constantly and highly expressed during the life cycle of the parasite^27,28^, while the acidic terminal sequence of the var genes (PfEMP1), detected by the PfvarATS assay, is associated with lower RNA levels^29^. The ability of the ENAR protocol to coextract DNA and RNA was also demonstrated with the following experiment: Five µL of an *in vitro-*generated stage V gametocyte culture was applied onto the RDTs and stored at RT for three weeks before NAs were extracted by ENAR. The gametocyte-specific transcript PF3D7_0630000 was reverse transcribed and amplified using a published assay which does not require DNase treatment for specific detection of gametocytes^30^. Extracted NAs from 5 μL undiluted and 1:100 diluted stage V gametocytes specifically amplified the gametocyte marker, while the control without a reverse transcription step did not result in amplification (Supplementary Figure S2).

The PfIS, an international standard with known parasite density, was used to explore the feasibility of quantifying *Pf* parasites extracted by ENAR. In total, 51 individual RDTs containing 5 µL PfIS with different parasite densities, ranging from 0.1 to 10,000 Pf/µL of the PfIS, were prepared. A high reproducibility and reverse correlation between parasite densities and Cq values were observed for both targets, the *Pf* specific PfvarATS and the pan-*Plasmodium* target Pspp18S (Fig 1D). Based on the slope, RT-qPCR efficiencies of 75.4% and 124.3% were calculated for PfvarATS and Psp-p18S, respectively. RDTs negative for PlasQ assay amplification (Cq > 45, colored in red) carried mostly dilutions representing parasite densities ≤ 1 parasite/µL. Two exceptions were observed where the Pspp18S assay failed to amplify two RDTs probed with higher parasitemia levels (5 and 10 Pf/µL, respectively). RDTs probed with 1 parasite/µL were detected in 4 (PfvarATS) and 7 (Pspp18S) out of 10 RDTs tested.

In summary, based on experiments conducted with standardized *Pf* reference samples we conclude that ENAR is able to recover both DNA and RNA, which results in an increased sensitivity of the RT-qPCR compared to the qPCR-based detection methods. The lower limit of detection (LOD) for RT-qPCR-based amplification of NAs from RDTs is around 1 Pf/µL, although 10x lower parasitemia levels can be detected as demonstrated with freshly cultured *Pf* parasites. RDTs are a reliable source of NAs and extraction by ENAR followed by analysis using RT-qPCR assays allows quantification of *Pf* parasites.

### Evaluation of ENAR protocol using Controlled Human Malaria Infection studies as a platform

Blood collected from volunteers undergoing Controlled Human Malaria Infection (CHMI) studies represent well-characterized samples as the parasite strain, the timing and dosing of infection is known. Therefore, blood samples collected from volunteers undergoing CHMI are well suited for developing and validating novel malaria diagnostic tools^31^.

The ENAR protocol was evaluated with venous blood samples collected during CHMIs assessing the efficacy of Sanaria’s PfSPZ Vaccine in clinical trials in Bagamoyo, Tanzania in malaria pre-exposed volunteers. RDTs were spiked with blood and stored as part of two CHMIs, the first of which was conducted in 2016 (CHMI-1) and the second in 2018 (CHMI-2). As part of the standard diagnostic procedures during the CHMIs, whole blood was collected in EDTA tubes and DNA extracted from a total of 180 µL whole blood. A DNA-based qPCR assay was run and parasitemia quantified (defined as WB-qPCR). Parasite densities as low as 0.05 Pf/µL are detected with the WB-qPCR protocol. During both CHMIs, fresh blood from asymptomatic subjects collected 9 to 18 days post-CHMI was tested with RDTs (Table 1). CHMI-1 and CHMI-2 used two different types of RDTs, which required 20 µL and 5 µL of whole blood, respectively. RDTs collected during CHMI-1 were stored for an average of 605 days (categorized as > 18 months), while RDTs collected during CHMI-2 were stored for an average of 18 days (< 1 month) before processing following the ENAR protocol. For the entire storage period, RDTs were kept at RT in a closed box and protected from light. NAs were extracted from the RDTs using the ENAR protocol and parasites were detected and quantified by RT-qPCR using the PlasQ assay.

**Table 1.** Overview of blood samples collected during two CHMIs and stored on RDTs.

| **Study (Year)** | **CHMI-1 (2016)**<br>**> 18 months storage** | **CHMI-2 (2018)**<br>**< 1 month storage** |
| --- | --- | --- |
| RDT brand | BinaxNOW® Malaria RDT | CareStart™ Malaria (Pf/PAN) Combo |
| Number of RDTs collected | 71 | 50 |
| Blood volume on RDT | 20 µL | 5 µL |
| Storage time in days<br>(mean and range) | 605 (596-616) | 18 (10-48) |
| Storage conditions | RT | RT |
| Sampling days post CHMI<br>(mean and range) | 14.0 (10.5-18.0) | 12.7 (9.0-18.0) |
| % positive by WB-qPCR | 38.0% (27/71) | 62.0% (31/50) |
| WB-qPCR parasite density<br>(parasites/µL, geom. mean and range) | 4.7 (0.05-840.0) | 0.3 (0.01-1041.0) |

### Impact of long-term storage on detection rate of parasite NA extracted by ENAR

First, we analyzed the impact of RDT storage time on parasite detection rates. Long-term storage (> 18 months) negatively affects the *Pf* detection rate in samples with a parasite density between 1 and 10 Pf/µL but has no negative impact on samples with initial parasite density greater than 10 Pf/µL (Fig 2A). Long-term storage negatively affects the detection rate based on the Pspp18S target (33% vs 100%, Fisher’s exact test p = 0.06) more than the PfvarATS target (66% vs 100%, Fisher’s exact test p = 0.46). Interestingly, the parasite densities estimated from RDTs with shorter storage time (< 1 month) are closer to the reference parasite densities assessed by WB-qPCR using 180 µL freshly prepared blood than the estimates from RDTs with longer storage time (> 18 months) (Fig 2B). This is an additional indicator that NAs conserved on RDTs might undergo degradation over time.

**Figure 2.**
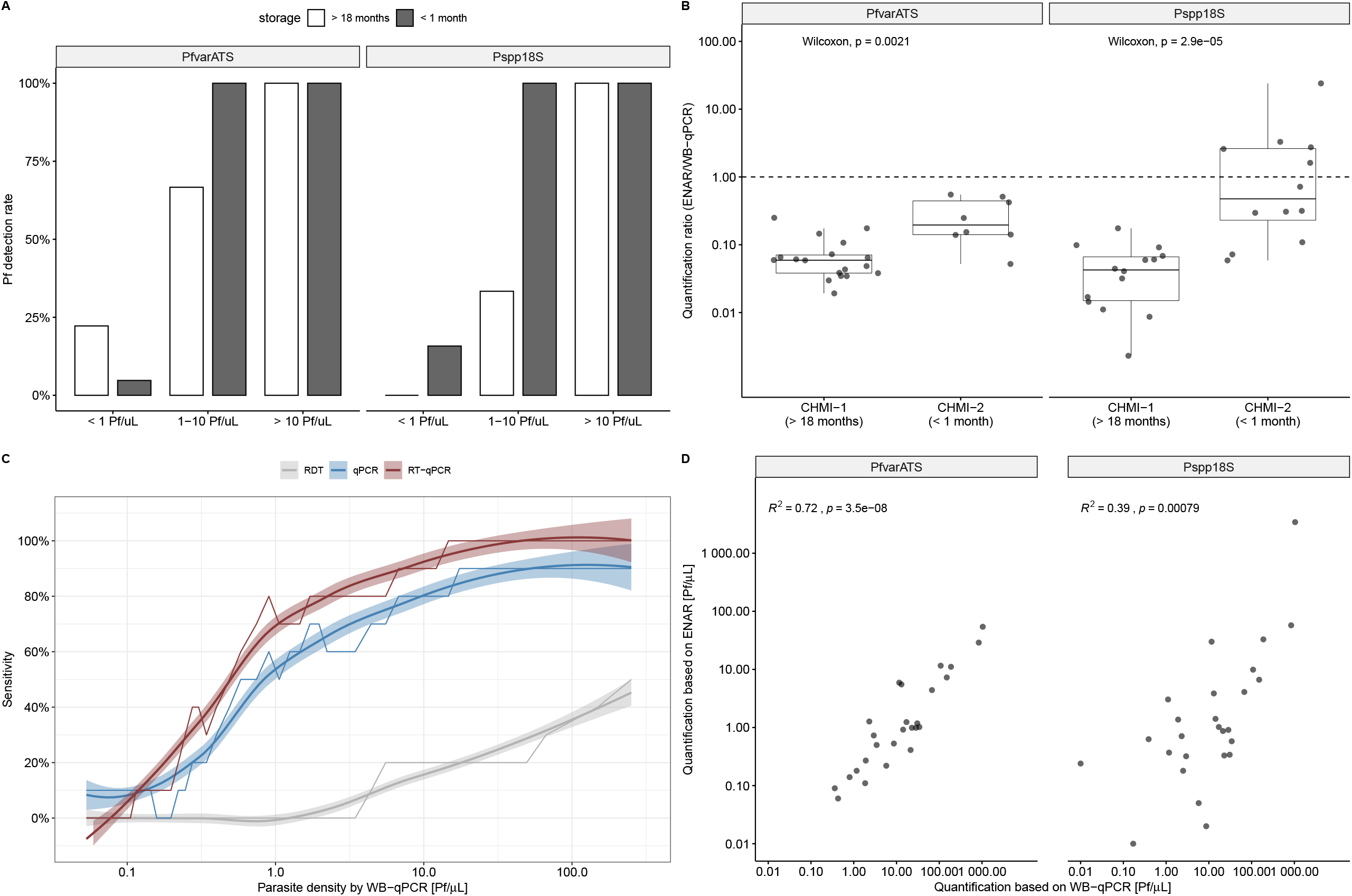
Evaluation of ENAR protocol using samples collected during CHMI studies. A) *Pf* detection rates grouped by parasite density and storage time. B) Quantification ratio between densities derived from ENAR and densities derived from whole blood qPCR (WB-qPCR). C) Diagnostic sensitivity of rapid diagnostic test (RDT), ENAR followed by qPCR detection (qPCR) and ENAR followed by RT-qPCR detection (RT-qPCR) in relation to parasite density. Rolling means of 10 observations, using WB-qPCR as a gold standard, are shown with 95% CIs (shaded areas). D) Correlation of parasite densities obtained from DNA extracted from fresh whole blood and NAs extracted by ENAR.

### Clinical sensitivity and parasite quantification based on ENAR approach

If the data of both CHMIs are combined, the overall detection rate was 54% for the ENAR-based RT-qPCR when compared to WB-qPCR, which was significantly higher than detection by microscopy (9%) or PfHRP2 antigen capture by RDT (12%) using the same samples.

In order to understand the contribution of RNA on the detection rates in this clinical sample set, we compared RT-qPCR with qPCR. Detection rates of RT-qPCR in relation to parasite density reveals an improved diagnostic performance over the whole range of *Pf* densities compared to qPCR (Fig 2C). RT-qPCR is significantly more sensitive than qPCR for the Pspp18S assay (27% vs 47%, Mc-Nemar test p=0.0026), but not for the PfvarATS assay (47% vs 47%, McNemar test p=1.0). Interestingly, among the long-term stored RDTs collected in 2016, the detection rate of the Pspp18S assay was also significantly higher for RT-qPCR compared to qPCR (52% vs 22%, McNemar test p =0.01). Even after long-term storage a significant proportion of (fragmented) RNA can be still extracted and used for RT-qPCR amplification.

Parasite densities determined by WB-qPCR versus densities obtained with the ENAR-based RT-qPCR method showed significant positive correlation supporting the quantitative character of our approach (Fig 2D). The correlation was stronger with the PfvarATS assay (r^2^ = 0.72) than with the Pspp18S assay (r^2^ = 0.39).

### Implementation of ENAR protocol within malaria indicator survey

We implemented the ENAR approach within a malaria indicator survey in which we aimed to screen for SNPs in the pfk13 propeller region to study the prevalence and type of mutations potentially associated with artemisinin resistance. We tested ENAR using samples and data derived from the 2018 BIMEP MIS which included more than 13,000 individuals (Fig 3A). Instead of disposing the RDTs after use, the tests were labeled with a barcode to connect each RDT with other survey data collected in questionnaires (Fig 3B). For each of these barcode-labeled RDTs, an extra informed consent for molecular analysis was obtained from the participants or their legal guardians. For the sorting and selection of distinct RDTs for analysis, we developed the *RDTselect* app (https://github.com/Sparclex/barcode-value-finder), a browser-based mobile phone application which identifies barcode-labeled RDTs based on an input list containing all barcodes of a certain selection (Fig 3C).

**Figure 3.**
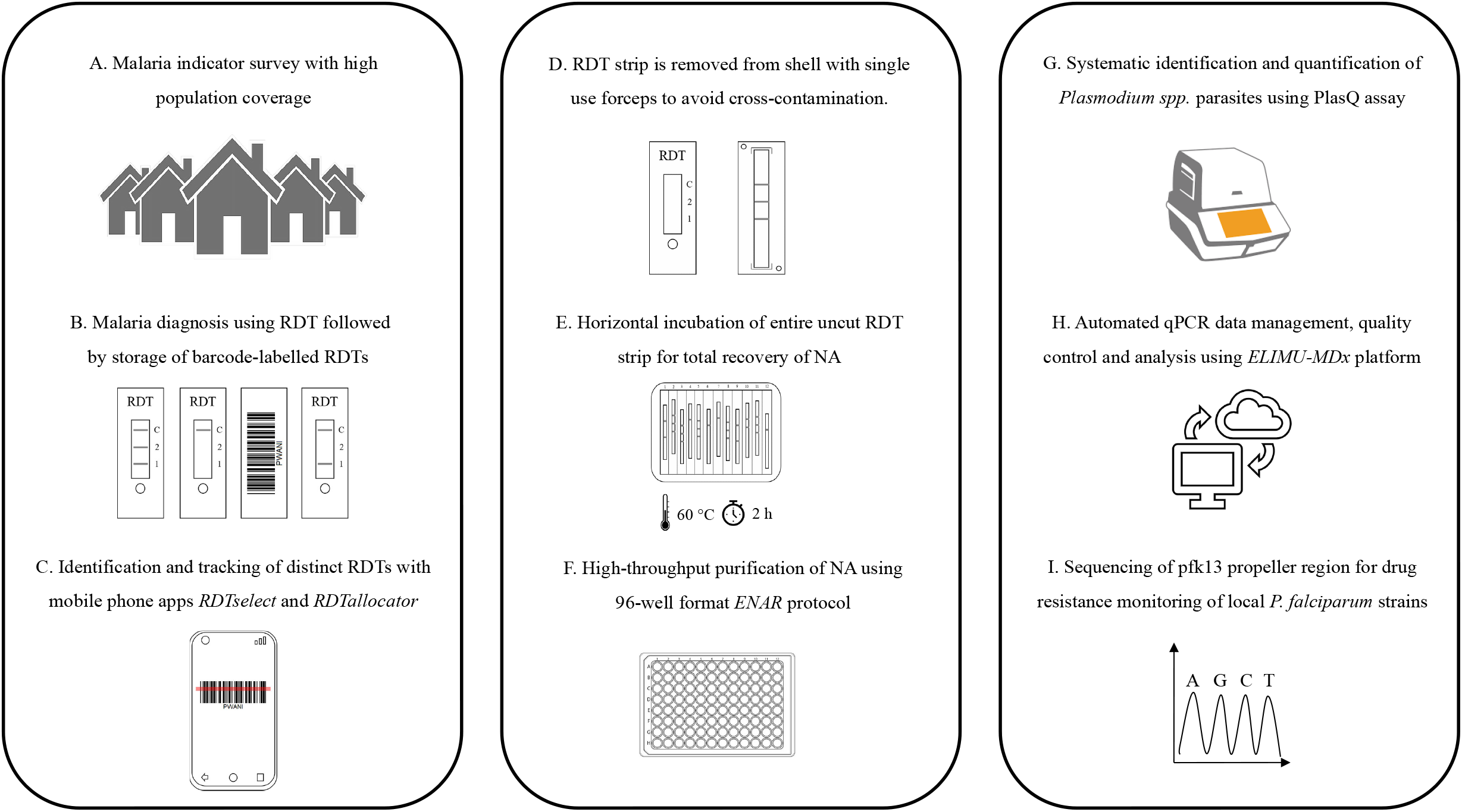
Adaptation of ENAR protocol for analyzing large numbers of barcoded RDTs. A) Malaria indicator survey conducted including a detailed questionnaire. B) Malaria prevalence is determined by RDT followed by storage of barcode-labelled RDTs. C) Sorting and tracking of RDTs using smartphone apps. D-F) High throughput protocol for extraction of NAs from RDTs using the ENAR approach. G) Detection and quantification of *Pf* and non-*Pf* malaria parasite. H) Automated analysis of qPCR data using ELIMU-MDx. I) Genotyping of pfk13 propeller region for drug resistance monitoring.

To enable tracking of an individual RDT throughout the ENAR extraction process the *RDTallocator* app (https://github.com/Sparclex/position-allocator) was programmed. The barcodes are scanned with a mobile phone camera and the *RDTallocator* app allocates the associated RDT strip to the next available position in a 96-well plate (Fig 3C). After opening the RDT shell the entire uncut RDT strip is removed with sterile, single-use forceps (Fig 3D), incubated with lysis buffer in a 12-well long-format plate (Fig 3E), and NAs are extracted in a high-throughput 96-well format of the ENAR protocol (Fig 3F). All extracted samples undergo initial screening for presence of *Plasmodium* spp. parasites and quality control using the PlasQ RT-qPCR assay (Fig 3G). All RT-qPCR data generated were managed and analyzed by a custom-designed laboratory management and information system. ELIMU-MDx is designed for automated quality control, management and analysis of qPCR data^32^ (Fig 3H). Samples positive for *Pf* were subjected to amplification and sequencing of pfk13 for identification of SNPs associated with drug resistance (Fig 3I).

A total of 2690 out of 13,270 (20.3%) RDTs were extracted by ENAR and analyzed for *Plasmodium* spp. parasites by RT-qPCR. The demographic information of the entire MIS population and the selected volunteers for the molecular analysis are given in Table 2. Only volunteers with body temperature < 37.5 °C were included. Volunteers with a positive RDT and pregnant women are intentionally over-represented in our sample set.

**Table 2.**
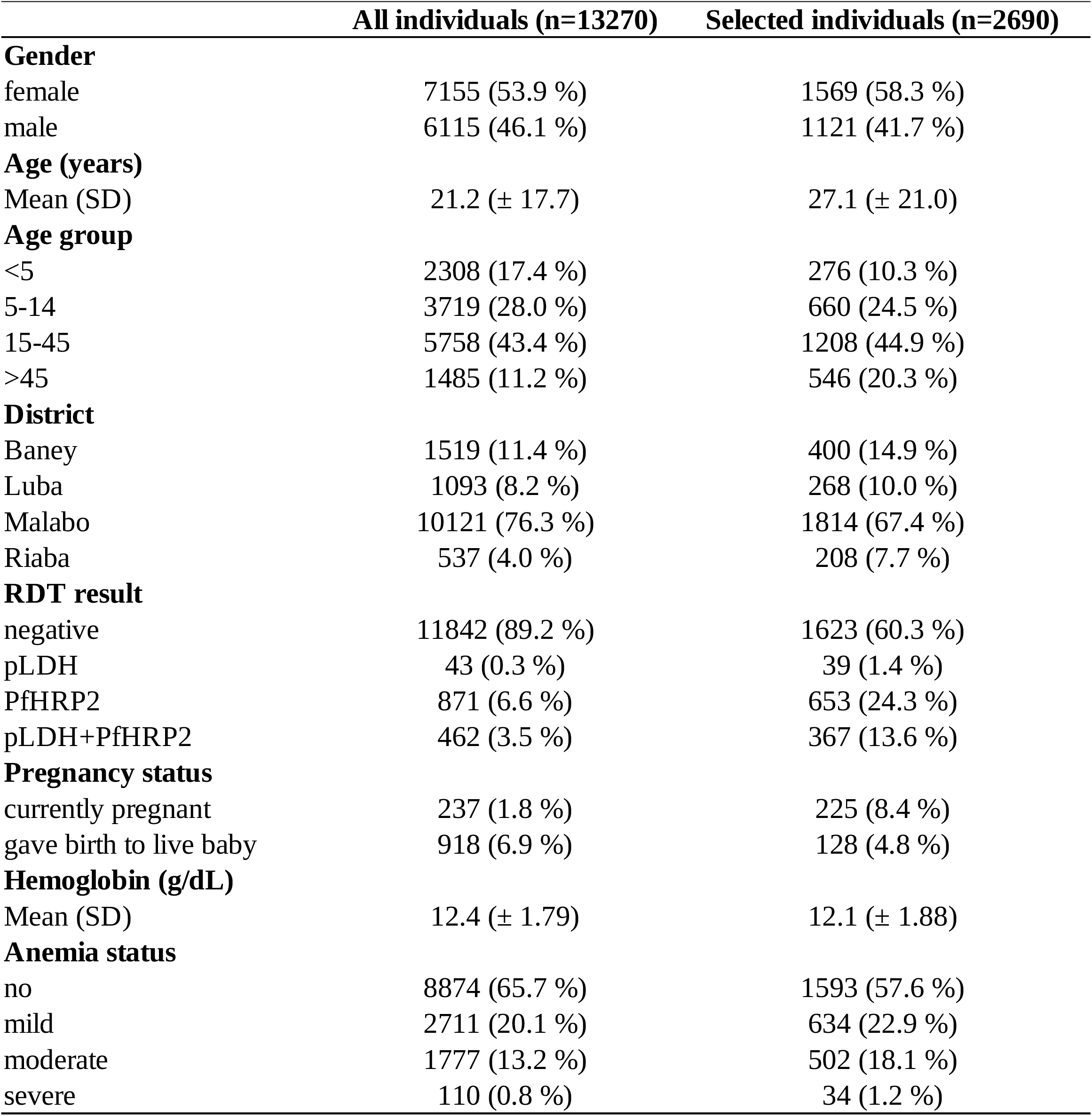
Demographic information of MIS participants.

### Malaria infections among asymptomatic MIS participants are characterized by *Pf* infections with mainly low parasite densities

Applying the approach described in Figure 3, 30.8% (828/2690) of the analyzed RDTs tested positive for *Plasmodium* spp. NAs (Table 3). A qPCR-based species identification revealed that 92.9% were *Pf*, 4.0% *P. malariae* and 1.0% *P. ovale* spp. No *P. vivax* or *P. knowlesi* NAs were found. In this asymptomatic population, *Pf* infections had on average parasite density of 29.2 Pf/µL, with densities being the highest among children below the age of five years (Fig 4a). The rather low parasitemia levels of asymptomatic individuals in combination with the small amount of blood available have implications for pfk13 genotyping. Samples with parasitemia levels below 50 Pf/µL are rarely amplified successfully for pfk13 sequencing (Fig 4b). In order to increase the efficiency of pfk13 genotyping process from RDTs, pre-selection based on RDT result is advised. For example, 84.5% of RDTs positive for both, PfHRP2 and pLDH carried parasite densities high enough to result in successful amplification of the pfk13 propeller region.

**Table 3.** ENAR-based identification of malaria parasites using PlasQ RT-qPCR assay.

|  | number of samples (%) |
| --- | --- |
| RDTs analysed by PlasQ | 2690 |
| Positive for PlasQ RT-qPCR | 828 ( 30.8% ) |
| ***Plasmodium* spp. identification** |  |
| Positive for *P. falciparum* | 769 ( 92.9% ) |
| *P. falciparum* with >100 Pf/μL | 227 ( 29.5% ) |
| Positive for *P. malariae* | 33 ( 4.0% ) |
| Positive for *P. ovale* spp. | 8 ( 1.0% ) |
| Positive for *P. knowlesi* | 0 ( 0.0% ) |
| Positive for *P. vivax* | 0 ( 0.0% ) |
| *Pf/Pm* co-infections | 16 (1.9%) |

**Figure 4.**
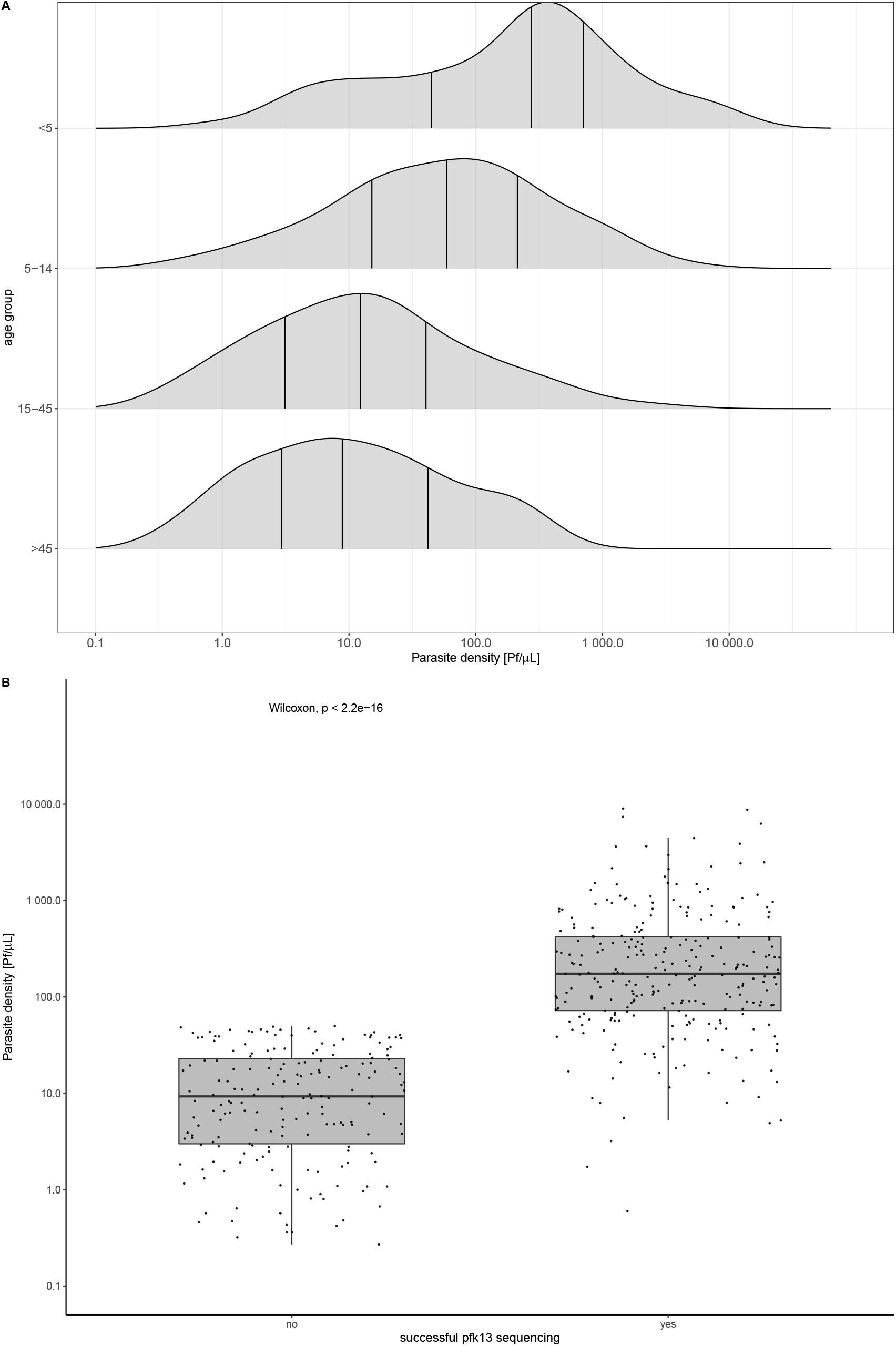
Parasite densities among asymptomatic individuals and implication for sequence analysis. A) Age group dependent parasite densities. B) Association between parasite density and successful amplification of pfk13 for sequence analysis.

### Low prevalence of SNPs in the pfk13 propeller region among *Pf* parasite isolates on Bioko

Sequence analysis of the pfk13 propeller region revealed a low prevalence of SNPs (Table 4). 97.6% (283/290) of Bioko’s *Pf* isolates carried the wildtype allele. Two isolates had the A578S and one the V589I non-synonymous SNP, which have been described in sub-Saharan Africa before^12,33^ and are not associated with artemisinin resistance^34^. The P553L SNP was first described in Cambodia^13^. This SNP has previously been found at low prevalence in East Africa, in Kenya and Malawi^35^ as well was recently found in an isolate from a Chinese national returned from Angola^36^. To our knowledge, the V517I SNP has never been described before. Compared to the other three known SNPs, the V517I SNP had the lowest PROVEAN^37^ score, indicating no or neutral effects on the biological function of the kelch 13 protein. Two synonymous SNPs, namely, V510V and C469C, were also found.

**Table 4.** *Pf*k13 propeller polymorphisms observed in MIS population on Bioko Island.

| **Kelch13 propeller genotyping** |  | **PROVEAN score** |
| --- | --- | --- |
| *P. falciparum* strains sequenced | 290 |  |
| PfNF54 allele | 283 ( 97.6% ) |  |
| **Non-synonymous SNPs** |  |  |
| A578S (G1732T) | 2 ( 0.69% ) | -1.962 |
| V589I (G1765A) | 1 ( 0.35% ) | -0.663 |
| V517I (G1549A) | 1 ( 0.35% ) | -0.562 |
| P553L (C1659T) | 1 ( 0.35% ) | -1.721 |
| **Synonymous SNPs** |  |  |
| V510V (G1530A) | 1 ( 0.35% ) |  |
| C469C (C1407T) | 1 ( 0.35% ) |  |

## Discussion

This report presents the development of an approach for large-scale, high-throughput and cost efficient molecular surveillance of malaria parasites based on extraction of NAs from RDTs. During the development of ENAR, special attention was given to the evaluation of its reproducibility and the impact of long-term storage on the detectability of the NAs. Using samples from CHMI studies as a standardized platform allowed us to conclude that NAs can be reliably recovered and amplified from RDTs, even after long-term storage at RT. The small amount of blood in combination with low parasite density is a challenge when it comes to detecting *Pf* in asymptomatic patients. Therefore, we aimed to maximize the amount of NA recovered from RDTs by expanding the pool of possible target molecules to RNA by using RT-qPCR. Even after a storage period of over 18 months at RT, the detection rate of the RT-qPCR assay was still significantly higher compared to qPCR only, indicating long-term preservation of DNA and RNA.

We aimed to transform the ENAR approach into an flexible tool for larger scale surveillance studies by increasing extraction and analysis throughput. The ENAR approach was successfully integrated into the 2018 BIMEP MIS on Bioko Island. More than 13,000 individuals gave extra consent for storage and molecular analysis of their RDT. This high acceptance rate was also described by others^21^ and can be attributed to the convenience of blood collection by finger prick and the small blood volume, usually 5 to 10 µL, needed for RDTs. With a total of 2750 RDTs, we analyzed blood from more than 20% of the MIS participants. This was made possible by the development of custom-made software solutions for sorting and identification of RDTs and by a significant reduction in processing time by using the entire RDT strip instead of cutting it into pieces.

Robust (quantitative) data, as generated by ENAR, in combination with a large-scale MIS adds substantial value to our understanding of malaria endemicity on Bioko Island without conducting additional expensive and time consuming epidemiological studies. In addition this process allows for researches to detect various species of malaria parasites. For instance, we found *P. malariae* and *P. ovale* spp., but did not find *P. vivax*, as in previous studies when surveys carried out in 1996 and 1998 found two^38^ and one^39^ case of *P. vivax* infection on Bioko Island.

In addition, we screened for SNPs in the propeller region of the pfk13 gene among asymptomatic individuals to obtain data of possible artemisinin-resistant *Pf* strains circulating on the island. We found that 1.7% (5/290) of the analyzed *Pf* isolates had non-synonymous SNPs in the pfk13 propeller region, which is comparable to the prevalence found in other African countries ^33^. Among the five isolates with non-synonymous SNPs, two isolates had the A578S, one the V589I, one the P553L and one the V517I SNP. The A578S and V589I allele had been reported in the region already^40,41^, and we found one new previously unreported non-synonymous SNP, V517I. Interestingly, the P553L SNP is the only mutation we found which was previously associated with delayed parasite clearance^12^. Although the prevalence of pfk13 SNPs seems to be low in the moment, the spread of *Pf* parasites with pfk13 SNPs needs to be closely monitored. A molecular surveillance approach as presented may offer a unique opportunity to support policy makers regarding choice and change of drugs for malaria treatment^42^.

Based on the presented results, we propose that ENAR provides a powerful tool for molecular malaria surveillance and could be reliably used for retrospective quantitative and in-depth molecular studies of malaria.

## Material and methods

### *Pf* reference samples

*Pf* reference samples were used to test the performance of the ENAR procedure. Experiments with *Pf* reference samples were conducted using Carestart^™^ HRP2/pLDH Combo RDTs (Access Bio, Inc., Somerset, NJ, USA). Serial dilutions of the WHO International Standard for *Pf* DNA Nucleic Acid Amplification Techniques (NIBSC code: 04/176, herein referred to as PfIS)^43^ were used to quantify *Pf* parasitemia by (RT)-qPCR. Whole blood was spiked with different parasite densities, ranging from 10,000 to 0.1 Pf/µL and 5 µL of this suspension applied onto RDT.

Additionally, ten-fold serial dilutions, ranging from 10,000 to 0.1 Pf/µL, of freshly cultured *Pf* strains PfNF54, PfDD2 and PfHB3 were prepared and 5 µL were applied onto RDTs. 5 µL of stage V gametocytes were obtained from *in vitro* parasite culture as described previously^44^. RDTs probed with these stage V gametocytes were extracted using the ENAR protocol after a three-week storage period at RT.

### School-based survey in Mkuranga district

Carestart^™^ HRP2/pLDH Combo RDTs were used to determine the parasite rate among asymptomatic children from three primary schools in the Mkuranga district of Coastal Tanzania. Extraction protocol A, which is based on the Quick-DNA^™^ Miniprep Kit (Zymo Research Corporation, Irvine CA, USA), was used to extract DNA from a total of 190 RDTs collected during this school-based survey. *Pf* was detected by amplifying the acidic terminal sequence of the var genes (PfvarATS)^45^.

### Sample collection, analysis and storage during CHMI studies

RDTs were collected during two CHMI studies conducted to evaluate Sanaria’s PfSPZ Vaccine in Bagamoyo, Tanzania (Clinical Trials.gov registration numbers NCT02613520 and NCT03420053, respectively). The first CHMI was conducted in 2016 (referred to CHMI-1) and the second CHMI was conducted in 2018 (referred to CHMI-2). Fresh venous whole blood collected in EDTA tubes was analyzed by RDTs within 45 min after blood collection. During CHMI-1, 20 µL was applied to BinaxNOW^®^ Malaria RDT (Alere, Cologne, Germany) and during CHMI-2, 5 µL was applied to Carestart^™^ HRP2/pLDH Combo RDT. The RDTs were read according to the manufacturers guidelines and then stored in a box at RT until extraction of NA.

The same samples were used to monitor parasitemia during CHMI by thick blood smear microscopy and qPCR as described elsewhere^46,47^. All samples were processed and analyzed at the laboratory of the Bagamoyo branch of the Ifakara Health Institute in Tanzania.

### Malaria indicator survey on Bioko Island, Equatorial Guinea

The 2018 BIMEP Malaria Indicator Survey (MIS) was carried out between August and October 2018 on a representative sample of 13,505 individuals from 4774 households selected from all communities across Bioko Island. All consenting permanent residents and short-term visitors were tested for malaria using the CareStart™ Malaria HRP2/pLDH Combo RDT. Used RDTs were stored at RT in plastic bags containing desiccants and transported to the Swiss Tropical and Public Health Institute for further molecular analysis.

### Nucleic acid extraction methods from RDTs

The RDT cassettes were opened, the entire RDT strip removed and cut into four small pieces in order to fit into a 1.5 mL micro-centrifuge tube. A set of cleaned forceps and scissors were used with special attention given to prevent cross-contamination between samples. After processing a sample, the scissors and forceps were cleaned in 10% sodium hypochlorite, wiped with ethanol-sprayed tissues and dried before processing the next sample. The four nucleic extraction protocols tested, named A through D, were all based on silica columns.

#### Protocol A – ZR Quick-DNA™ Miniprep Kit

The protocol is based on the Quick-DNA™ Miniprep Kit (Zymo Research Corporation, Irvine CA, USA). Briefly, 1 mL of Genomic Lysis Buffer was added to the pre-cut RDT strip in a 1.5 mL micro-centrifuge tube and incubated at 95 °C for 20 minutes. The mixture was then transferred onto the extraction column and the manufacturers guide was followed for extraction. DNA was eluted in 50 µL of DNA Elution Buffer.

#### Protocol B – ZR Quick-DNA™ Miniprep Plus Kit

The protocol is based on the Quick-DNA™ Miniprep Plus Kit (Zymo Research Corporation, Irvine CA, USA). We added 600 μL of Solid Tissue Buffer (Blue) and 40 μL of Proteinase K to the pre-cut RDT strip in a 1.5mL micro-centrifuge tube and incubated at 55 °C for 60 minutes. The supernatant was transferred to a clean 1.5 mL micro-centrifuge tube and 640 μL of Genomic Lysis Buffer was added and thoroughly mixed. The mixture was transferred onto the extraction column and extracted per manufacturers guidelines. DNA was eluted in 50 µL of DNA Elution Buffer.

#### Protocol C – NukEx Pure RNA/DNA Kit

The protocol is based on NukEx Pure RNA/DNA Kit (Gerbion GmbH, Kornwestheim, Germany), which co-extracts DNA and RNA. We created a working solution of 500 μL of Binding Buffer, 4 μL of Poly A and 50 μL of Proteinase K. The working soution was added to the pre-cut RDT strip in a 1.5 mL micro-centrifuge tube following incubation at 60 °C for 10 minutes. The supernatant was transferred onto the NukEx Spin Column and textraction was carried out per manufacturer’s guidelines. Total NAs were eluted in 50 µL of Elution Buffer.

#### Protocol D – Zainabadi et al. extraction method for DBS

The protocol is based on a recently published extraction protocol for total NAs from dried blood spots^48^. Identical buffer compositions were used, and the protocol was adapted to extraction of NAs from RDT strips. We incubated the pre-cut RDT strip in 900 µL lysis buffer at 60 °C for 2 hours. The supernatant was transferred onto Omega HiBind RNA Mini Columns (Omega Bio-Tek, Norcross, USA) and NAs extracted as described. NAs were eluted in 50 µL of Elution Buffer (Quick-DNA™ Miniprep Kit, Zymo Research Corporation, Irvine CA, USA).

### High-throughput extraction protocol of NAs from RDTs (ENAR protocol)

We adapted protocol D to extract NAs from used RDTs in a high-throughput manner. The main modification included a horizontal incubation of the entire uncut RDT strip by using sterile, RNase-/DNase-free 12-channel reservoirs (Axygen, Corning Inc, USA) and switching to a 96-well format for extraction. By removing the cutting step, the hands-on time during the extraction process is significantly reduced, as well the risk of cross-contamination by carryover during the cutting process is minimized. Up to eight 12-channel reservoirs, with a total of 96 samples, were processed in one batch. Lysis was conducted by adding 900 µL lysis buffer to each RDT strip placed in the 12channel reservoir followed by incubation at 60 °C for 2 hours with gentle shaking. All supernatants were then transferred to Omega E-Z 96 wells DNA plates (Omega Bio-Tek, Norcross, USA), washed with Wash Buffer 1 and 2 and lastly eluted into a 96 well plate (DNA LoBind Plates, Eppendorf) with 50 µL pre-warmed (60 °C) Elution Buffer (Zymo Research Corporation, Irvine CA, USA). With these adaptations to the protocol, NA from 96 RDTs can be extracted in about three hours, with minimal hands-on time needed. One positive control (RDT spiked with 5 µL blood containing 200 Pf/µL) and one negative control (Lysis Buffer only) were included with each extraction plate to control for plate-to-plate consistency and cross-contamination. A standard operating procedure (SOP) for ENAR can be found in Supplementary Protocol S1.

### Detection and quantification of *Plasmodium* spp. parasites

We used the PlasQ assay, a multiplex qPCR assay for *Plasmodium* spp. and *Pf* detection and quantification to analyze the NAs extracted from RDTs^47^. The PlasQ assay targets the Pan-*Plasmodium* 18S DNA and RNA (Pspp18S)^49,50^ and the *Pf*-specific acidic terminal sequence of the var genes (Pf-varATS)^45^. The human *rnasep* gene (HsRNaseP)^49^ served as an internal control to assess the quality of NA extraction and qPCR amplification. To run the PlasQ as a RT-qPCR assay, targeting both DNA and RNA templates, we added 1x Luna WarmStart RT Enzyme Mix (New England Biolabs, Ipswich, USA) and started the RT-qPCR program with a reverse transcription step at 55 °C for 15 min. All qPCR and RT-qPCR assays were run on a Bio-Rad CFX96 Real-Time PCR System (BioRad Laboratories, California, USA). Samples were analyzed in duplicate with positive (PfNF54 DNA), negative (malaria negative individual) and non-template (molecular biology grade H_2_O) controls added to each qPCR run.

### Multiplex pre-amplification of *Plasmodium* spp. DNA

The *Plasmodium* spp. 18S rDNA and pfk13 genes of all PlasQ-positive samples were amplified in a multiplex reaction by conventional PCR. We amplified 3 μL of extracted NAs in a total volume of 20 μL using 1x HOT FIREPol® MultiPlex Mix (Solis Biodyne, Tartu, Estonia). Using 0.25 µM of the published primers, AGT GGA AGA CAT CAT GTA ACC AG and CCA AGC TGC CAT TCA TTT GT, 986 bp of the pfk13 propeller region were amplified^26^. Simultaneously, 1407-1469 bp of the pan-*Plasmodium* 18S rDNA were amplified using 0.5 µM of GRA ACT SSS AAC GGC TCA TT^51^ and AGC AGG TTA AGA TCT CGT TCG^49^. The conditions of the multiplex PCR were the following: 95 °C for 12 minutes; 25 cycles of 95 °C for 20 seconds, 57 °C for 40 seconds and 72 °C for 1 minute 45 seconds; and 72 °C for 10 minutes.

### Detection of gametocytes and *Plasmodium* spp. species identification

#### Gametocyte-specific RT-qPCR assay

A previously published RT-qPCR assay for identification of *Pf* gametocytes based the PF3D7_0630000 transcript was used^30^. Briefly, 2 µL of extracted NAs were added to 8 µL reaction mix consisting of 0.6 µM of primers, 0.3 µM probe and Luna® Universal Probe One-Step RT-qPCR Kit (New England Biolabs, Ipswich, USA). The qPCR program included a reverse transcription step for 15 min at 53 °C, followed by polymerase activation for 1 min at 95 °C, and 45 cycles with 15 s at 95 °C and 45 s at 53 °C.

#### Plasmodium spp. species identification

Non-*falciparum Plasmodium* species identification based on the 18S rDNA gene was performed. 2 µL of the product from the *Plasmodium* spp. multiplex pre-amplification were added to the master mix containing 1x Luna® Universal Probe qPCR Master Mix, 0.8 µM forward (GTT AAG GGA GTG AAG ACG ATC AGA) and 0.8 µM reverse primers (AAC CCA AAG ACT TTG ATT TCT CAT AA) to amplify a 157-to 165-bp segment of the *Plasmodium* spp. 18S rDNA gene^52^. Species-specific probes were selected to differentiate between the species. *P. malariae* was detected using a Yakima Yellow-labelled MGB probe (CTA TCT AAA AGA AAC ACT CAT) ^53^, *P. ovale* spp. using a novel designed Texas Red-labelled and LNA-modified probe (GGA [LNA-A]AT [LNA-T]TC TTA GAT TGC TTC CT[LNA-T] CAG), *P. vivax a* Cy5-labelled probe (GAA TTT TCT CTT CGG AGT TTA)^54^ and *P. knowlesi* a Cy5-labelled probe (CTC TCC GGA GAT TAG AAC TCT TAG ATT GCT)^55^. The conditions for the qPCR were: 95 °C for 3 minutes and 45 cycles of 95 °C for 15 seconds and 57 °C for 45 seconds.

### Genotyping of pfk13 propeller region

In a second PCR reaction with a 15 μL total volume, 1.5 μL of the product from the *Plasmodium* spp. multiplex pre-amplification was amplified using 1x HOT FIREPol® MultiPlex Mix (Solis Biodyne, Tartu, Estonia) and 0.33 μM forward (TGA AGC CTT GTT GAA AGA AGC A) and reverse (TCG CCA TTT TCT CCT CCT GT) primers. Except for an annealing temperature of 58 °C, the PCR conditions were similar to the first reaction. The 798 bp product of the second PCR was evaluated using agarose gel electrophoresis and samples which failed amplification were repeated. Amplicons were sequenced by Microsynth (Microsynth AG, Balgach, Switzerland).

### Data analysis and statistics

All (RT)-qPCR assays were run in duplicates and initial data analysis of the (RT)-qPCR data was conducted using CFX Maestro Software (Bio-Rad Laboratories, California, USA). In the case where one replicate interpreted as positive and the other negative, then the assay was repeated and the result was considered positive if two positive replicates were obtained out of the total four replicates. All (RT)-qPCR data generated were managed and analyzed by a custom-designed laboratory management and information system named ELIMU-MDx^32^. The ELIMU-MDx platform supports automated quality control, management and analysis of qPCR data. Oligo design and sequence analysis was performed using Geneious Prime 2019.1.1 (https://www.geneious.com). Statistical analysis and visualization of data was conducted using R version 3.5.1 based on packages *dplyr, ggpubr, ggplot2, gridextra, reshape2* and *scales*.

## Data Availability

The datasets supporting the conclusions of this article are included within the
article.

## Acknowledgments

Etienne Guirou and Charlene Yoboue are recipients of Swiss Government Excellence Scholarships (Number 2016.1250 and 2017.0748, respectively) granted by the State Secretariat for Education, Research and Innovation. We would like to thank Christin Gumpp, Christian Scheurer and Sergio Wittlin from the Swiss TPH Malaria Drug Discovery Group for their help with cultivating PfNF54, PfDD2 and PfHB3 parasites. We are grateful to Eva Hitz and Till Voss from the Swiss TPH Malaria Gene Regulation Unit for kindly providing with *Pf* gametocytes culture.

## Authors’ contributions

Conceptualization: EAG, TS, CD

Data curation and validation: EAG, TS, OTD

Formal analysis and visualization: EAG, TS

Funding acquisition: CD, MT, CM, BMN

Investigation: JS, NS, HM

Methodology: EAG, SH, GC, AD, LG, MM, CAY

Resources: SA, NS, JS, SLH, GM, CCF, WPP, GAG

Software: SK

Project administration and supervision: CD, TS

Writing – original draft: EAG, TS, CD

## Ethics approval and consent to participate

For the school-based survey in Mkuranga district, sample collection was approved by the Senate Research and Publication Committee (SRPC) of the Muhimbili University of Health and Allied Sciences and the respective authorities at Mkuranga district.

Both clinical trials were performed in accordance with Good Clinical Practices (GCP). CHMI-1 (Clinical Trials.gov: NCT02613520) protocol was approved by IRBs of the Ifakara Health Institute (IHI) (Ref. No. IHI/IRB/No: 32-2015), the National Institute for Medical Research Tanzania (NIMR) (NIMR/HQ/R.8a/Vol.IX/2049), and the Ethikkommission Nordwestund Zentralschweiz (EKNZ) Switzerland (reference number 15/104). The protocol was also approved by the Tanzania Food and Drug Authority (TFDA) (Auth. No. TZ15CT013). CHMI-2 (Clinical Trials.gov: NCT03420053) protocol was approved by IHI’s IRB (Ref. No. IHI/IRB/No: 32-2015), NIMR (NIMR/HQ/R.8a/Vol.IX/2049), EKNZ (reference number 15/104) and TFDA (Auth. No. TZ15CT013). The 2018 malaria indicator survey was approved by the Ministry of Health and Social Welfare of Equatorial Guinea and the Ethics Committee of the London School of Hygiene & Tropical Medicine. Written informed consent was obtained from all adults and from parents or guardians of children who agreed to participate. Only samples for which an additional consent for molecular analysis was obtained were included in this study.

We confirm that all experiments were performed in accordance with relevant guidelines and regulations.

## Abbreviations

*Pf*: *P. falciparum*
pfk13: *Pf* kelch 13
RDT: rapid diagnostic test
DBS: dried blood spot
ENAR: extraction of nucleic acids from RDT
CHMI: controlled human malaria infection
NA: nucleic acid
NAT: nucleic acid amplification technique
PfIS: WHO International standard for *P. falciparum* NAT
LOD: limit of detection
RT: room temperature
qPCR: quantitative polymerase chain reaction
PlasQ: multiplex qPCR assay for quantification of *P. falciparum* and *Plasmodium spp*. parasites
SNP: single nucleotide polymorphism
Pf/µL: *Pf* parasites per µL blood

## Supplementary Information

**Supplementary Table S1**. Summary of published studies using DNA extracted from RDTs for molecular analysis of malaria parasites.

**Supplementary Protocol S1**. Extraction of Nucleic Acids from RDTs (ENAR): step-by-step protocol

**Supplementary Figure S1**. Accumulation of captured *Pf* NAs on RDTs.

**Supplementary Figure S2**. Detection of the gametocyte-specific transcript PF3D7_0630000 in blood on RDTs after three weeks of storage at RT.

